# The Role of the BMPR2 pathway in Group-II Pulmonary Hypertension Secondary to Valve Heart Disease

**DOI:** 10.1101/2025.02.26.25321265

**Authors:** Jorge Martínez-Solano, Jaime Bermejo-Fernández, Ana González-Mansilla, Rocío García-Orta, Pedro L Sánchez-Fernández, Mario Castaño, Javier Segovia-Cubero, Pilar Escribano-Subías, Antoni Bayés-Genís, Pablo Martínez-Legazpi, Teresa Mombiela, Carlos Ortíz-Bautista, Arantxa González, Javier Bermejo, Ana I Fernández-Avila

## Abstract

BMPR2 is widely known to be key in the pathobiology of pulmonary arterial hypertension. Recently, data regarding pulmonary hypertension (PH) secondary to left-heart disease (LHD) have focused on inflammatory and proliferative pathways rather than classic hemodynamic scope but BMPR2 role is yet to be determined. In the current study, we assessed BMPR2 and its regulatory factor, SRSF2, in patients with PH due to valvular heart disease (VHD). Blood samples from patients included in the Sildenafil for Improving Outcomes after Valvular Correction (SIOVAC) clinical trial were analysed. The study involved PH-VHD patients with combined pre-and postcapillary PH, isolated postcapillary PH, control patients without PH and healthy subjects. We sequenced the BMPR2 exons, measured BMPR2 A-isoform expression and analysed SRSF2 gene expression levels in blood samples. Our results demonstrate that BMPR2 A-isoform levels are reduced in patients with PH-VHD compared to controls and, even more, healthy subjects. Furthermore, BMPR2 A-isoform expression showed meaningful prognostic value among PH-VHD patients. Notably, whereas pathogenic gene variants were ruled out, SRSF2 was indeed associated with BMPR2 A-isoform differential expression. To the best of our knowledge, this is the first study demonstrating the involvement of BMPR2 gene expression in PH-LHD patients, supports BMPR2 A-isoform as a potential prognosis biomarker and this pathway as a therapeutic target.

---

Pulmonary hypertension (PH) due to left heart disease (PH-LHD) represents a major health issue worldwide, accounting for nearly 75% of cases of PH. PH-LHD eventually develops in more than half of patients with heart failure and confers poor prognosis. It is recognized that targeted therapies are lacking partially because the molecular mechanisms underlying PH-LHD remain poorly understood [1,2].

The *Bone Morphogenetic Protein Receptor Type 2 (BMPR2)* gene plays a central role in pulmonary arterial hypertension (PAH), whose downregulation leads to the upregulation of the activin pathway regulated by ActRIIA, entailing cellular proliferation in PAH [3]. Human *BMPR2* produces two primary transcripts, the isoform-A, containing all 13 exons, is the functional protein. Notably, even in the absence of a heritable mutation, levels of the BMPR2 A-isoform are reduced in lung tissue and blood from patients with PAH [4]. Furthermore, the *Serine and Arginine Rich Splicing Factor 2* (*SRSF2*) regulates the balance between BMPR2 isoforms [5]. Although a link between *BMPR2* isoforms in PH-LHD has been suggested [3,6–8], this connection has not been stablished in the clinical setting. This study aims to evaluate the role of *BMPR2* and *SRSF2* in patients with PH due to valvular heart disease (VHD) and explore their potential as prognostic biomarkers.

We have conducted a *post-hoc* analysis of the Sildenafil for Improving Outcomes after Valvular Correction (SIOVAC) clinical trial [9], we included a cohort of adult patients in a stable clinical condition with a mean pulmonary artery pressure ≥□30□mmHg who had previously undergone a successful valvular correction procedure. After completing the 6-month treatment with sildenafil vs. placebo, patients underwent an extended follow-up. In addition, we studied groups of I) control patients with previous heart valve disease in whom we excluded PH after successful valve procedure, and 2) healthy subjects without cardiac disease.

First, we analyzed *BMPR2* gene variants in 109 patients (45 of them with combined pre and postcapillary PH, 42 with isolated postcapillary PH, and 22 controls). The 13 exons of the *BMPR2* gene were amplified and sequenced by Sanger from DNA samples. The analysis of *BMPR2* sequence data revealed a total of 12 variants in 47 of the 109 patients. *BMPR2* variants were present in 33% of patients with PH-LHD and 32% of control subjects. Based on ACMG criteria, only 5 PH-LHD patients (4.4%) carried a pathogenic variant (NM_001204.7[BMPR2]: c.1748dup[p.Asn583fs]).

Second, we analyzed gene expression in a subset of 76 subjects with available blood samples stored in Tempus Blood RNA Tubes for whole RNA isolation (49 PH-LHD patients, 19 controls and 8 healthy subjects). *BMPR2* A-isoform expression was measured by RT-qPCR using the primer pair: Fw-GTTCCAAGCACAGCAGCAGA and Rv-ACATTTCACAGACAGTTCATTCCT, designed on exon12-exon13 (**Fig. 1A**). The Kaplan–Meier method was used to characterize the global survival time conditioned on *BMPR2* expression using a median threshold. In addition, we measured the expression of the *SRSF2* gene using the primer pair Fw-CGACGCTGAGGACGCTATGGATGC and Rv-CGGCTGCGAGACCTGGAACGACT. *GADPH* as control gene.

**Fig. 1.**
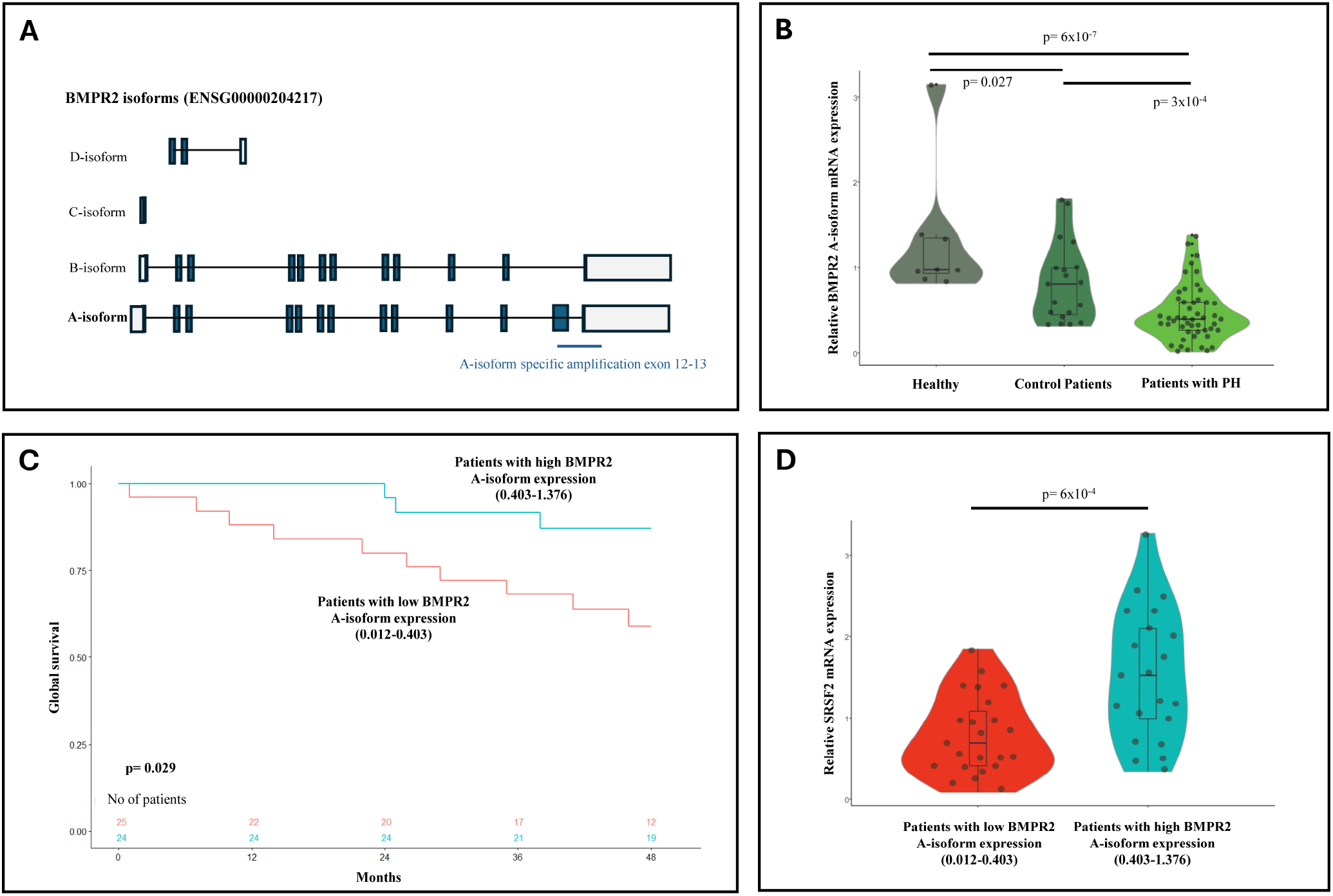
*BMPR2* A-isoform gene expression in PH-VHD patients. (A) *BMPR2* isoforms. (B) Expression differences in healthy subjects, control patients and patients with PH after a successful valve procedure. (C) Kaplan–Meier analysis for global survival at 4 years. (D) *SRSF2* expression differences in PH-VHD patients with low and high levels of *BMPR2* A-isoform expression

Expression of the *BMPR2* A-isoform varied significantly across the three groups (**Fig. 1B**). Patients with PH-LHD exhibited 1.8-fold lower expression in A-isoform levels compared to controls and 2.8-fold lower expression compared to healthy subjects. Notably, patients with PH-LHD and low BMPR2 A-isoform expression (< median value) had reduced 4-year survival compared to those with high expression (≥ median; **Fig. 1C**).

Additionally, patients with low BMPR2 A-isoform levels also showed lower levels of *SRSF2* splicing factor (**Fig. 1D**). Baseline clinical characteristics, including age, sex and cardiovascular risk factors, and pulmonary vascular resistance (combined vs isolated post-capillary PH) showed no association with the expression of *BMPR2* or *SRSF2*.

To the best of our knowledge, this is the first study to establish the involvement of *BMPR2* gene expression in patients with group-II PH. Herein we demonstrate that expression of the *BMPR2* A-isoform is significantly reduced in patients with PH-VHD, particularly among those with poorer 4-year survival outcomes. Pathogenic gene variants were excluded as the cause of this differential expression; instead, the variability in *BMPR2* A-isoform expression was related to differences in the *SRSF2* splicing regulator.

A certain degree of overlap between PAH and PH-LHD has been proposed, as they share common mechanisms and histopathological features [1]. However, the lack of efficacy of pulmonary arterial vasodilators in all forms of group-II PH suggests the presence of distinct molecular mechanisms between both conditions [7,10]. Importantly, currently available pulmonary vasodilators target downstream metabolic pathways and do not influence *BMPR2* levels. The imbalance between *BMPR2* and activin-class ligands plays a crucial role in PAH, and as reported in animal studies, may also contribute to the development of PH-LHD and heart failure [6–8]. In this context, our study provides indirect evidence, for the first time in patients, supporting a potential role of activin inhibitors in PH-LHD, as currently being investigated (NCT04945460). Further research is needed to clarify the molecular and inflammation pathways involved in the postcapillary remodeling changes that drive PH-LHD [1,2,7,10].

In summary, our findings highlight the involvement of *BMPR2* isoform expression in the pathogenesis of PH-LHD, suggest its potential as a novel prognosis biomarker, and support the development of new therapeutic approaches targeting this BMPR2-ActRIIA pathway.

## Data Availability

All data produced in the present study are available upon reasonable request to the authors

## Acknowledgements

We are in debt to all the investigators involved in the SIOVAC clinical trial for their assistance in sampling and recording patient data. We thank Ana Fernández Baza for her assistance in the logistics of the study.

## Declarations

All authors have reported that they have no relationships relevant to the contents of this paper to disclose.

## Ethical Approval

The study received approval of Spanish Agency for Drugs and Medical Products, Spanish Ministry of Health (EC07-90772). All patients signed the informed consent for the study. A second approval by the ethics committees was obtained for the purpose of extending follow_Jup, which waived the need for signing a new informed consent (Comité de Ética de la Investigación con Medicamentos del Hospital General Universitario Gregorio Marañón, Madrid, Spain (CEIm 13/19).

